# Perceived racial discrimination and eating habits: Systematic review and conceptual models

**DOI:** 10.1101/2021.08.04.21261615

**Authors:** Ylana Elias Rodrigues, Marcos Fanton, Ricardo Suñé Novossat, Raquel Canuto

**Author notes:** Correspondence: Raquel Canuto. Post-graduate Program in Food, Nutrition and Health, Federal University of Rio Grande do Sul, UFRGS. Rua Ramiro Barcelos 2400, Rio Branco. 90035-003. Porto Alegre, RS, Brazil. This project received financial support from the Brazilian National Council for Scientific and Technological Development (CNPq, Process n. 420342/2018-4 and 439073/2018-9). Conflict of interest: None. This project was registered in PROSPERO (CRD42019142605).

## Abstract

**Context:** there is no systematic organization of the available evidence about the relationship between racial discrimination and eating habits. Also, there is no consensus about its causal mechanism.

**Objectives:** a systematic review was conducted to synthesize evidence on the association between perceived racial discrimination and eating habits (eating behavior and food consumption). A conceptual model was developed to depict the most common mechanisms used to explain this association.

**Data Sources:** articles and dissertations were retrieved from the PubMed, EMBASE, Lilacs, and PsycINFO databases from inception to January 2021.

**Data Analysis:** 19 studies were included. The assessment of methodological quality was conducted using the Newcastle-Ottawa Scale. On average, the studies showed a methodological quality of 66%. Forty-six associations were evaluated. There were 38 associations between perceived racial discrimination and negative eating habits, 29 concerning eating behavior, and 17 regarding food consumption.

**Conclusions:** perceived racial discrimination negatively affects eating habits. A broader conceptual framework based on ecosocial theory is suggested to guide future research that would include different racial discrimination dimensions, such as internalized and structural.

## Introduction

Previous epidemiological studies have shown the association between racial discrimination and health outcomes in specific populations, such as Black, Asian, and Indigenous individuals ^1, 2^. Most of these studies have a focus on interpersonal racial discrimination. This phenomenon is mainly investigated by perceived racial discrimination (PRD) measures and evaluates chronic stress due to unfair treatment experiences over victims’ lifetime ^3, 4^. As a chronic stressor, racial discrimination leads to psychological stress and affects health

behaviors, such as eating behaviors, physical activity, alcohol consumption, and smoking ^1, 5^. Thus, experiences accumulated over the lifetime harms physical health ultimately driving to non-communicable chronic diseases (NCD), such as hypertension ^6, 7^, heart conditions and illnesses ^2^, diabetes ^8^, obesity and overweight-related measures ^9–11^. Causal mechanisms used to explain these associations are based on different stress pathways, such as physiological, psychological, and behavioral ^5^.

Eating habits are the main factors involved in NCD etiology and its complex concept can be evaluated in different ways, including food consumption and eating behavior. Food consumption represents food and nutrients intake per se and eating behavior involves the psychological and cultural aspects of eating and its practices ^12^. Several studies have demonstrated an association between racial discrimination experiences throughout life and poor eating habits in the last decade ^4, 13–15^. In 2020, a systematic review of the impact of psychological factors (stress, anxiety, depression, and discrimination) on emotional eating and weight among American Black women suggested that negatively perceived stress may be predictive of emotional eating ^15^. As a recent manuscript published in the American Journal of Clinical Nutritional recommended, researchers should consider more seriously the impact of race and the effects of racism and racial discrimination in nutritional outcomes. Thus, they advocate the use of ethnicity as a more comprehensive framework to address health disparities that goes beyond biological markers and individual behaviors ^16^.

Despite the growing body of evidence on this issue, there is no systematization of the available evidence about the relationship between racial discrimination and eating habits, including food consumption and eating behaviors, from worldwide studies. Also, no consensus is established about its causal mechanism, remaining unclear how the several hypotheses and pathways developed by the primary studies can be integrated. Thus, we conducted a systematic review to examine the association between PRD and eating habits and analyzed the association’s pathways presented in the primary studies. We also synthesized the main pathways through a conceptual model and proposed a new and more complex one, with variables and constructs from social epidemiological theories.

## Methods

Preferred Reporting Items for Systematic Reviews and Meta-Analyses (PRISMA) was used to perform this study, which was registered in PROSPERO (CRD42019142605). The PICO (Population, Intervention/Exposure, Comparison, and Outcome) strategy was used to construct the research question (Table 1). Thus, the studies retrieved from the literature met the following inclusion criteria: (i) the study design was observational; (ii) the article derived from original research; (iii) population study consisted of adolescents, adults, and elderly from any racial group (iv) the exposure measurement included any quantitative measure of self-reported perception of racial discrimination, within all time frames; (v) comparison group (not exposed) included the groups presenting lower levels of perceived racial discrimination than the exposure group or those not exposed to racial discrimination; (vi) the outcome was eating habits, including food and beverage consumption, dietary patterns, dietary quality, and eating behavior (overeating, emotional eating, and dietary restraint). Studies that measured eating disorder pathologies were not included.

**Table 1.** The PICO (Population, Intervention, Comparison, Outcome) criteria

| **Criteria** | **Description** |
| --- | --- |
| Population | Adolescents, adults, and elderly from any racial group. |
| Intervention/Exposure | Quantitative measure of self-reported perception of racial discrimination, within all time frames. |
| Comparison | Groups presenting lower levels of perceived racial discrimination than the exposure group or those not exposed to racial discrimination. |
| Outcome | Eating habits, including food and beverage consumption, dietary patterns, dietary quality, and eating behavior (overeating, emotional eating, and dietary restraint). |

### Search strategy

The search was based on articles, dissertations, and theses retrieved from the PubMed, EMBASE, Lilacs, and PsycINFO databases published up to September 2020. The search was updated in January 2020, and no new publications were found. The terms were defined according to the Medical Subject Headings (MeSH) Terms and the keywords used in similar studies. The search strategy used in the Pubmed was: (Racism OR Interpersonal discrimination OR Perceived discrimination OR Perceived racial discrimination OR Perceived racism OR Racial/ethnic disparities OR Self-reported discrimination OR Self-reported racism OR Unfair treatment) AND (Diet Western OR Fast Foods OR Eating OR Fruit OR Healthy Diet OR Vegetables OR Dietary Fats OR Feeding Behavior OR Diet Surveys OR Diet quality OR Diet OR Dietary behavior OR Food consumption OR Food pattern OR Sweets OR hyperphagia OR Food Addiction OR Food Fussiness OR Emotional eating OR Dietary restraint OR Binge eating OR Loss of control eating) NOT (Feeding and Eating Disorders). No filters were used in the searches. Also, reference lists of the selected articles and publications of the research groups on discrimination and health were screened.

### Study selection

Two reviewers (YR and RN) independently conducted the review process. Initially, search results were imported into Mendeley and duplicate deleted. The reviewers read all titles and abstracts in order to assess eligibility for inclusion. Even studies with titles with broad outcomes, such as health and well-being, were included since eating habits could be a covariate that appears only in the full text. Afterward, the reviewers read the full text of each study remaining to assist screening for final inclusion.

### Data extraction and risk of bias assessment

Data were extracted into an Excel spreadsheet independently by the two reviewers, including (i) study design (ii) authors and year of publication (iii) population (iv) sample characteristics (v) age (vi) a description of the exposure measure (vii) description of tools/instruments to measure the exposure, number of items and exposure time frame (viii) a description of the outcome measure (ix) description of the tools/instruments to measure the outcome, number of items and exposure time frame (x) explanatory variables (xi) results (measures of association between exposure and outcome). In case of discrepancies between reviewers, a third reviewer (RC) was called upon to decide.

Newcastle-Ottawa Quality Assessment Scale (NOS) was used to assess each study’s risk of bias and quality. This scale was created initially to evaluate cohort and case-control studies using a star system ^17^. We adopted a modified version also used to assess cross-sectional studies ^18^. The instrument consists of three constructs: selection, comparability (or exposure in cross-sectional studies), and outcome. The maximum score is 8 for the cohort study and 10 for the cross-sectional study design. Higher scores indicate a higher study’s quality. In the present study, the number of stars was transformed into a quality percentage to compare studies regardless of design. This instrument allows the evaluation of the methodological quality of observational studies and is easily applicable. The two reviewers independently conducted the quality assessment process. After that, results were compared, and a consensus was reached.

### Qualitative analysis

The results were interpreted by their different characteristics and association measures, which vary between studies, using the narrative synthesis method. The results will be presented according to the outcome group (food consumption or eating behavior). The associations of perceived racial discrimination and eating habits were examined according to the study’s characteristics. Meta-analysis was not performed due to the heterogeneity of outcomes in the primary studies.

We also analyzed the theoretical approaches and causal mechanisms presented in the studies to explain the associations between exposure and outcomes. As Nancy Krieger has convincingly argued, along with different works, theory matters ^19–21^. Epidemiological frameworks are theories of disease distributions and, explicit or implicit, shape how we generate useful scientific knowledge and what kind of concepts and causal determinants we judge as testable and relevant. They integrate data gathering, scientific constructs, causal mechanisms, and biological and social pathways. They also help us understand who is accountable for the different and inequitable disease patterns in our societies. Thus, to make these epidemiological frameworks more explicit, we developed conceptual models to organize and synthesize the main mechanisms the studies used to test their hypothesis ^22, 23^. Finally, we discuss and propose a new and more complex conceptual model based on the ecosocial framework to guide future researches on this topic.

## Results

### Study selection

The flowchart of the article selection procedure is presented in Figure 1. The initial search resulted in 3576 articles. After excluding duplicates, 2453 articles were screened through titles and abstracts; at this stage, 2933 were excluded. The full text of 3 potential studies was not available. We tried to reach the authors, but these theses did not present contact information, or after contact attempted, they did not answer. We expected the answer for six months. Remained 54 articles to read thoroughly and assess eligibility. After reading, 35 studies were removed, as they did not meet the inclusion criteria. Ultimately, 19 studies were included in this review. Reference lists of each study were screened, but any new studies meeting inclusion criteria were identified.

**Figure 1.**
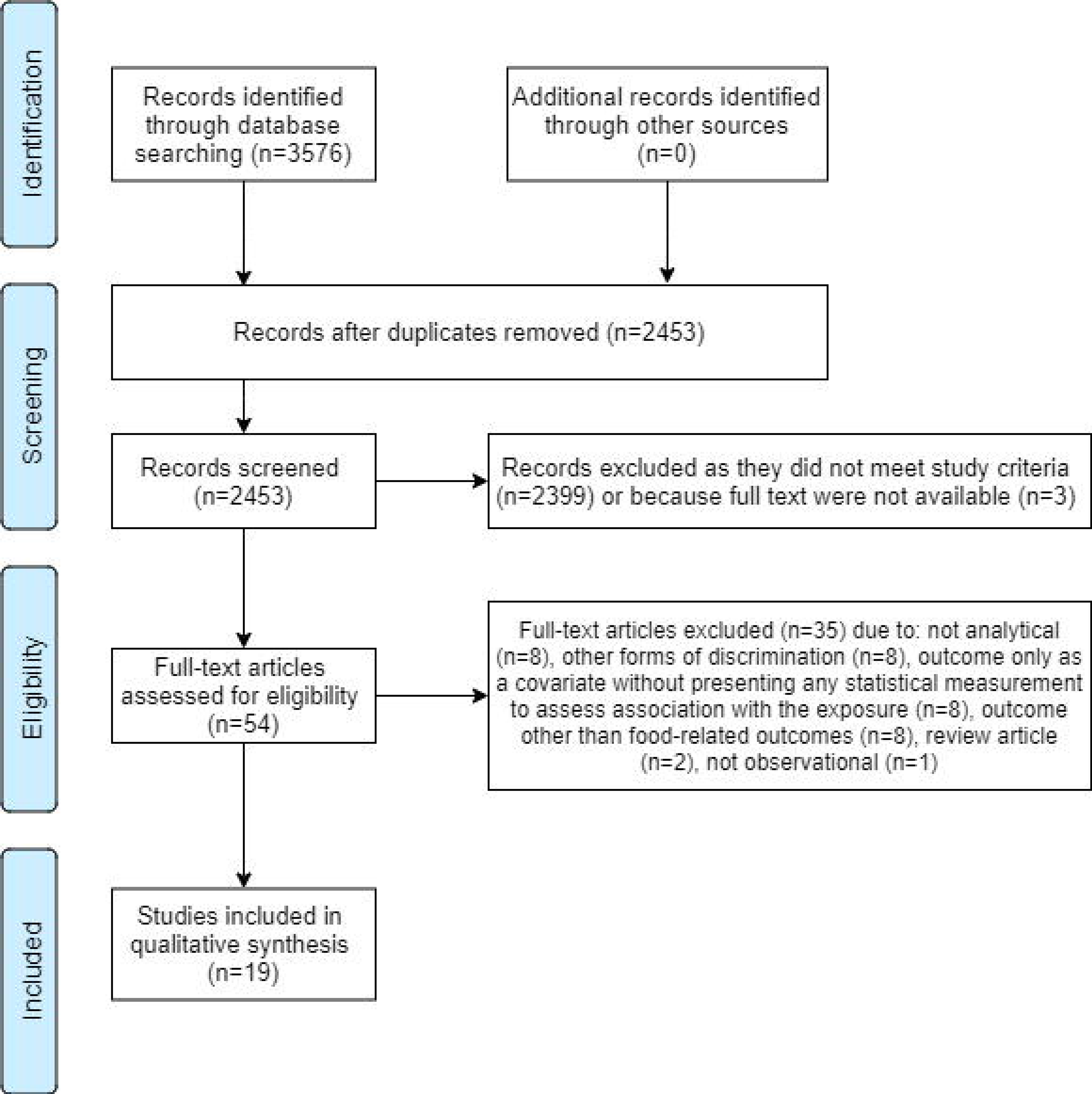
Flowchart of the systematic review on the association between Perceived Racial Discrimination (PRD) and eating habits.

### Study characteristics

Table 2 presents the general characteristics of the studies included in the review. All studies used a US-based population dataset. The year of publication ranged from 2004 to 2020, with 58% published between 2014 and 2020. Of the 19 studies included, only 4 were prospective cohorts ^13, 24–26^. The sample size ranged from 149 to 4925 subjects; 53% included less than 499 subjects.

**Table 2.** General characteristics of studies included in the review (**N=19)**.

| **Variables** | **N** | **%** |
| --- | --- | --- |
| **Country** |  |  |
| United States | 19 | 100 |
| **Study design** |  |  |
| Cross-sectional | 15 | 79 |
| Prospective | 4 | 21 |
| **Date of publication** |  |  |
| 2004-2008 | 2 | 11 |
| 2009-2013 | 6 | 32 |
| 2014-2020 | 11 | 58 |
| **Sample size** |  |  |
| 149-499 | 10 | 53 |
| 500-999 | 6 | 32 |
| 1000-4925 | 3 | 16 |
| **Race/ethnicity** |  |  |
| Black or African-American only | 12 | 63 |
| Black and White | 2 | 11 |
| Ethnic minorities | 2 | 11 |
| Native Americans only | 1 | 5 |
| South Asians only | 1 | 5 |
| Latinxs only | 1 | 5 |
| **Gender** |  |  |
| Both | 12 | 63 |
| Female only | 4 | 21 |
| Male only | 2 | 11 |
| All | 1 | 5 |
| **Lifecycle\*** |  |  |
| Adults | 19 | 100 |
| Elderly | 6 | 32 |
| Undergraduate students | 5 | 26 |
| Children | 1 | 5 |
| **Methodological Quality Assessment (%)** |  |  |
| 30-60% | 8 | 42 |
| 70-80% | 8 | 42 |
| 85%< | 3 | 16 |

\*Some articles had more than one category within the sample

Participants’ race/ethnicity in the majority of the studies (63%) focused on the Black or African-American population only ^4, 13, 14, 24–30, 31, 32^. The majority (63%) of the studies included female and male gender ^8, 13, 24–30, 32, 33, 34^. Some studies (32%) included the elderly ^14, 27, 28, 32–34^.

### Risk of bias within studies

Of the 19 studies, 8 scored between 30-60% ^8, 13, 14, 25, 26, 29, 30, 35^; 8 presented 70-80% ^4, 28, 32–34, 36–38^ and only 3 scored above 85% ^24, 27, 31^ (Table 2). The average percentage was 66%. The 4 cohorts reached 44% ^13^, 50% ^25^, 87,5% ^24^ and 50% ^26^.

Detailed information on the risk of bias is available in the supplementary material (S1). Since eating habits evaluation requires self-report information, it does not allow objective measurement of the study’s outcome, such as an independent blind assessment or record linkage. The NOS scores self-report assessment less than others, affecting the overall quality rating. Also, in cohorts, the NOS does not score for self-reported evaluation of the outcomes, decreasing the quality score.

### Perceived Racial Discrimination

Table 3 presents characteristics related to the exposure and outcomes of the studies included in this review. Among the studies that used validated instruments to measure PRD ^4,8,32–38,13,14,25–27,29–31,^ 26% used The Schedule of Racist Events ^25, 26, 30, 33, 35^ and 21% used Everyday Discrimination Scale (EDS) ^27, 34, 36, 38^. The others (42%) used a variety of validated scales ^4, 8, 13, 14, 29, 31, 32, 36^. Two studies used non-validated questionnaires ^24, 28^. Regarding the time frame, 42% of the studies focused on PRD experienced in the past year ^8, 26–29, 34, 36, 38^. Other studies (32%) used lifetime discrimination only ^13, 14, 24, 32, 33, 37^. Only 2 studies combined past year and lifetime discrimination ^4, 25^. Most of them measured the frequency of racial discrimination experienced in a variety of settings. Only 2 studies ^24, 36^ focused on specific settings (workplace or college and PRD following postpresidential elections).

**Table 3.**
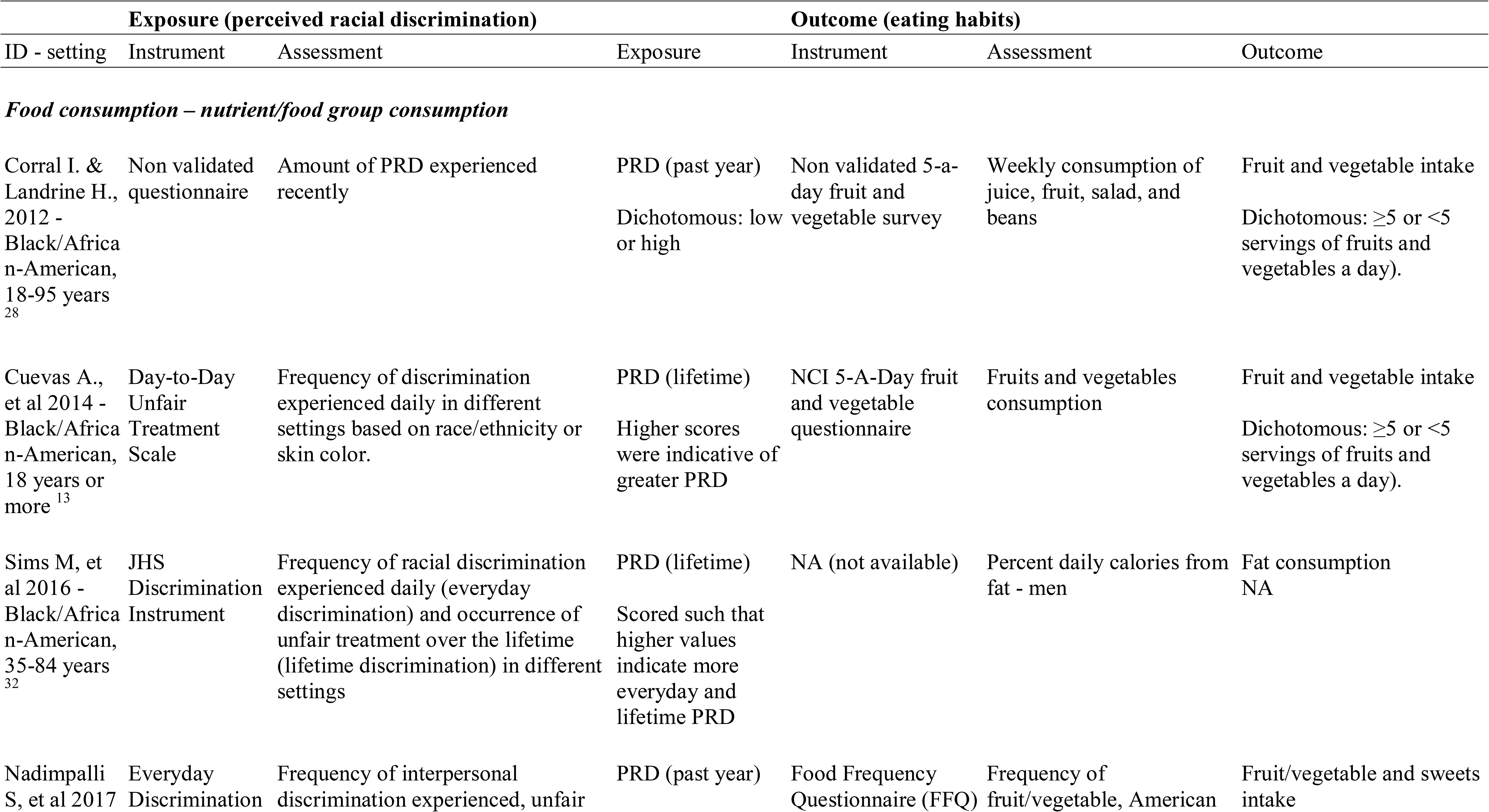
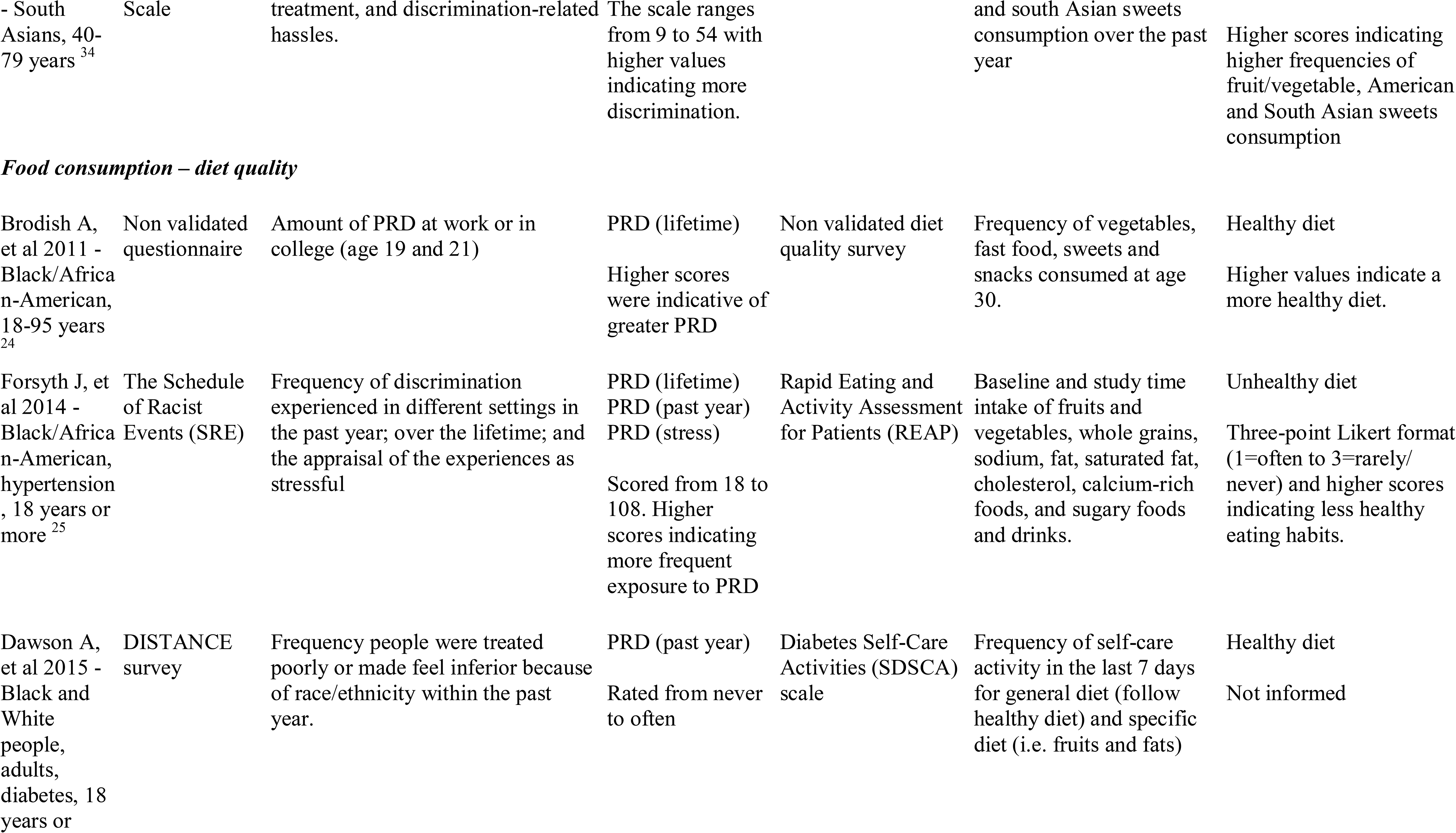
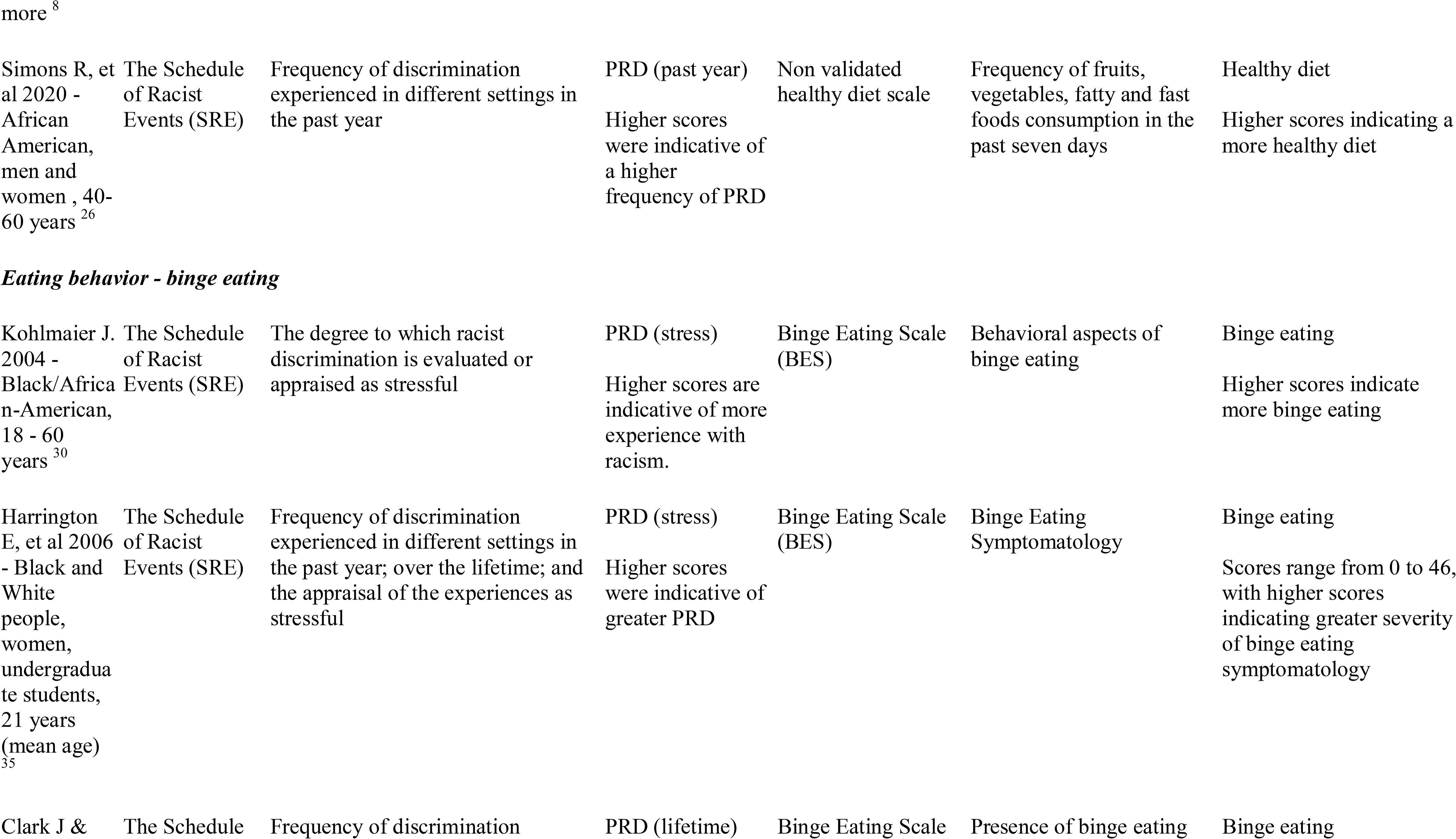
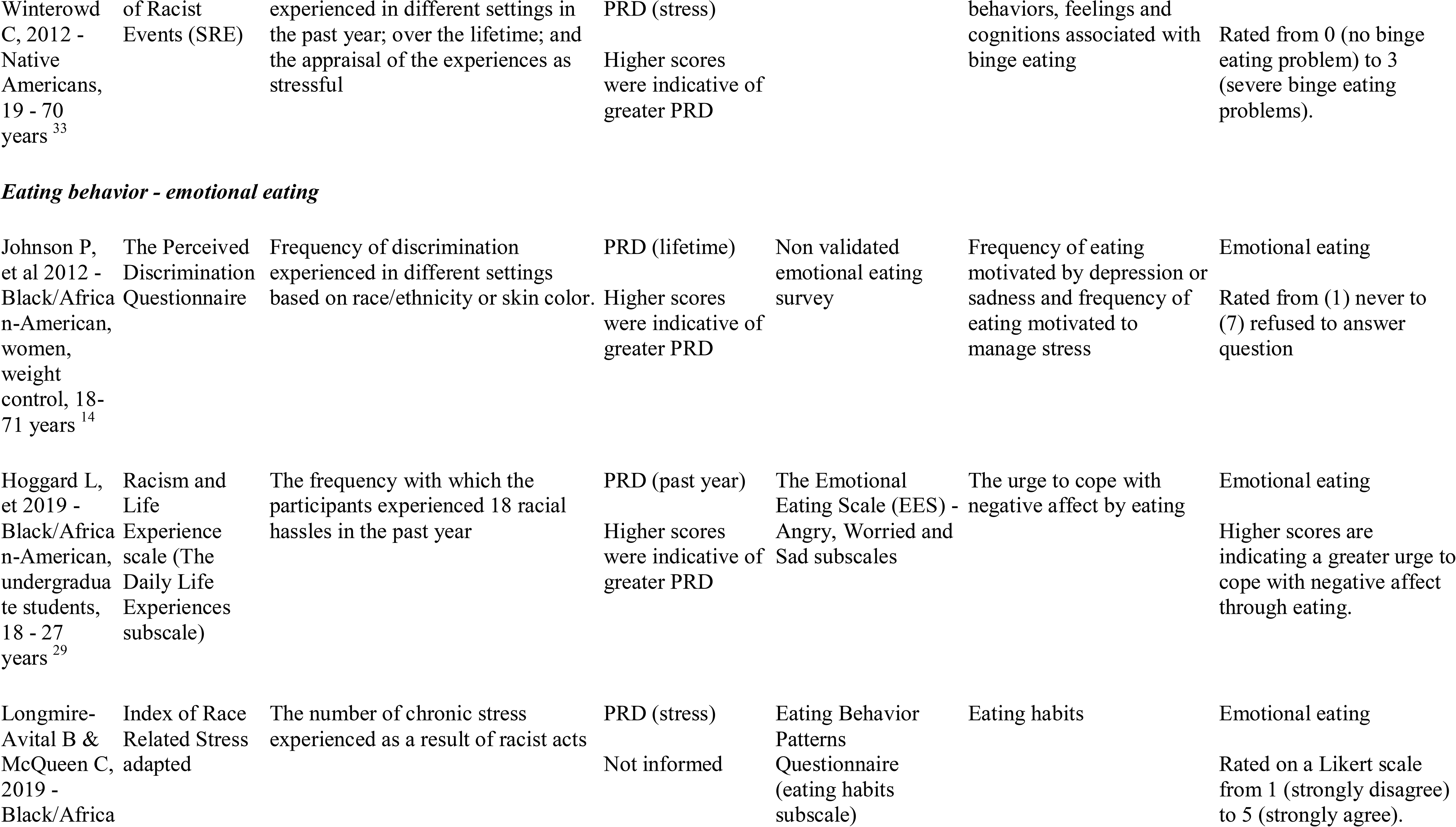
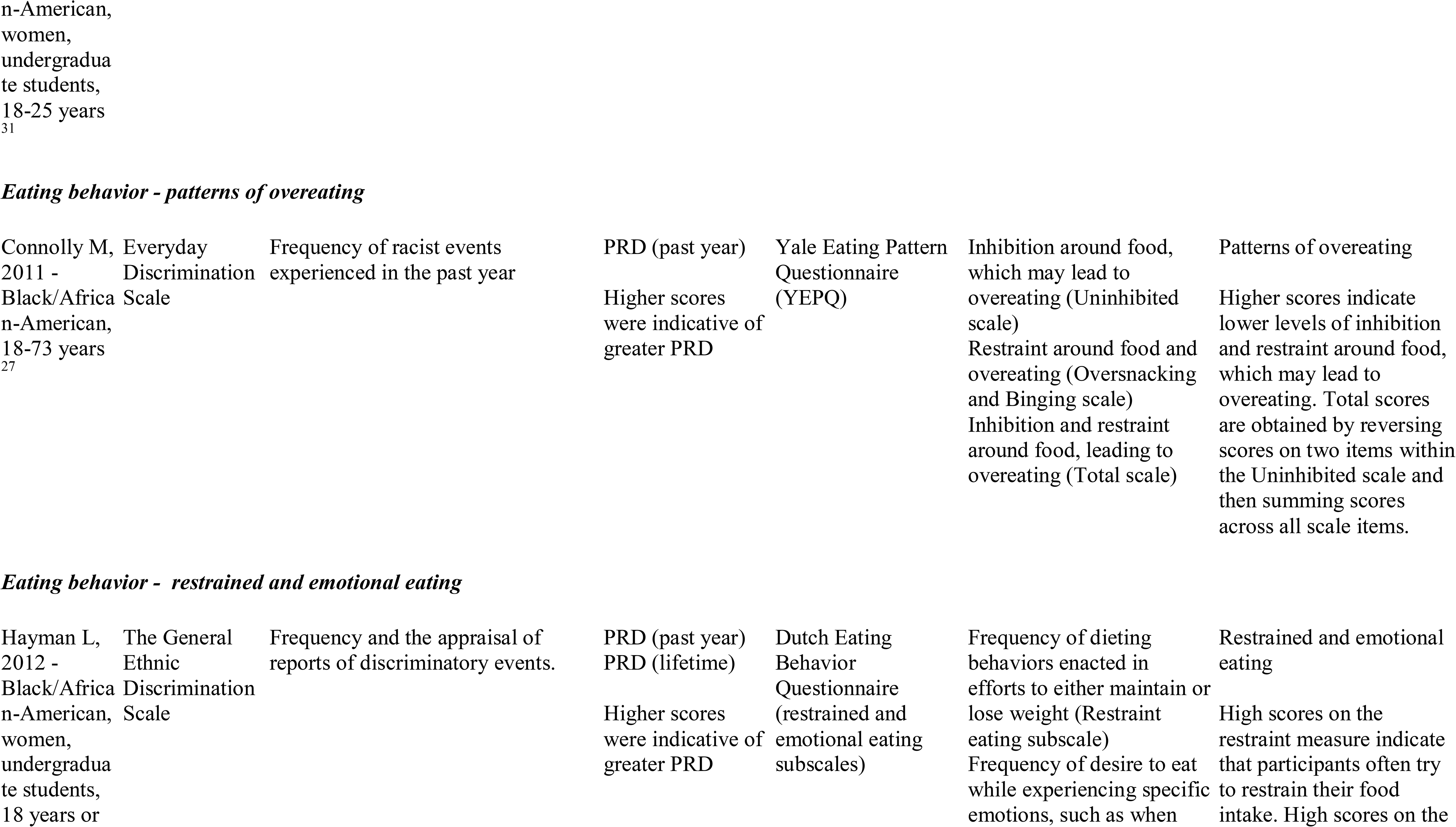
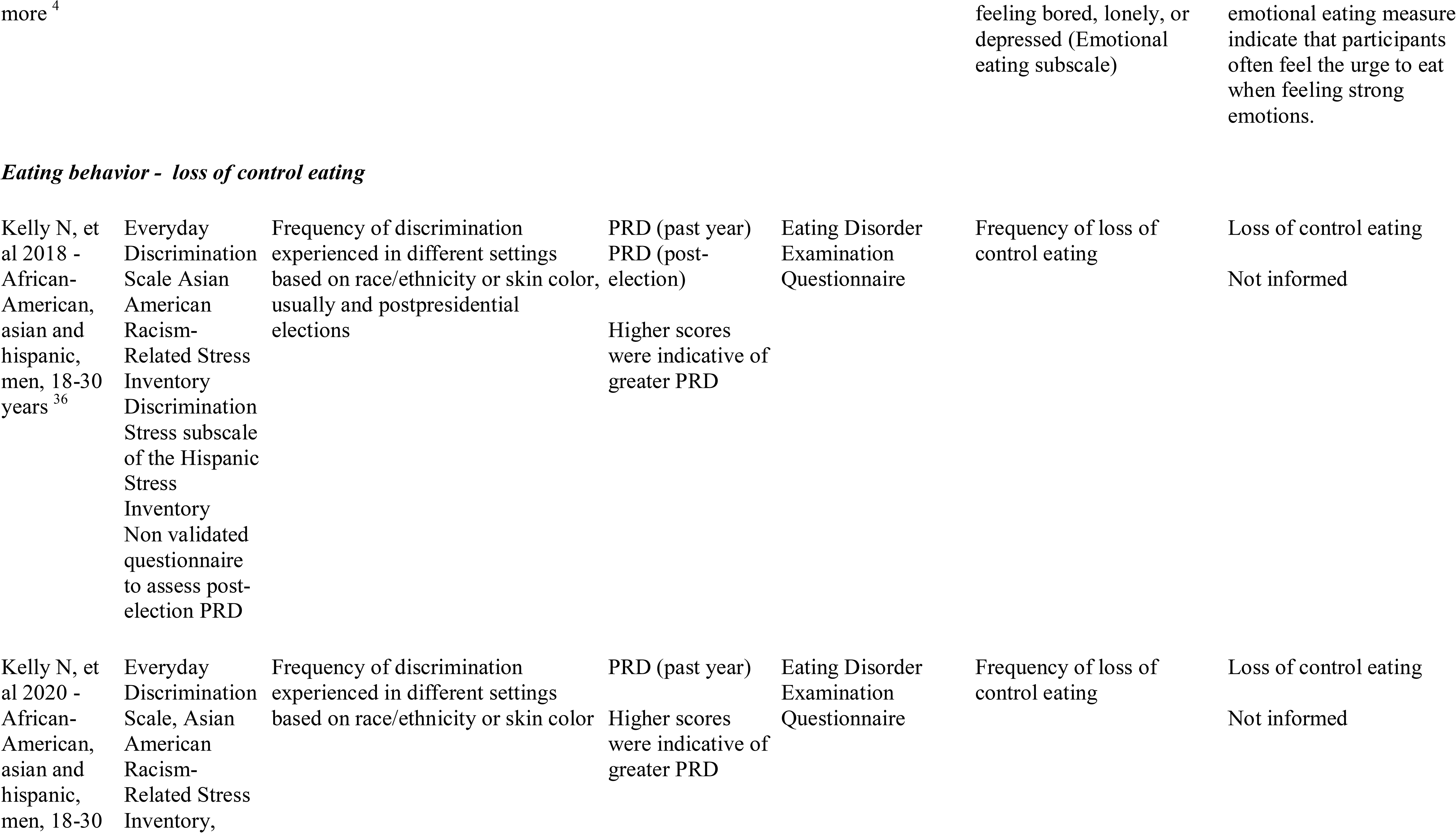
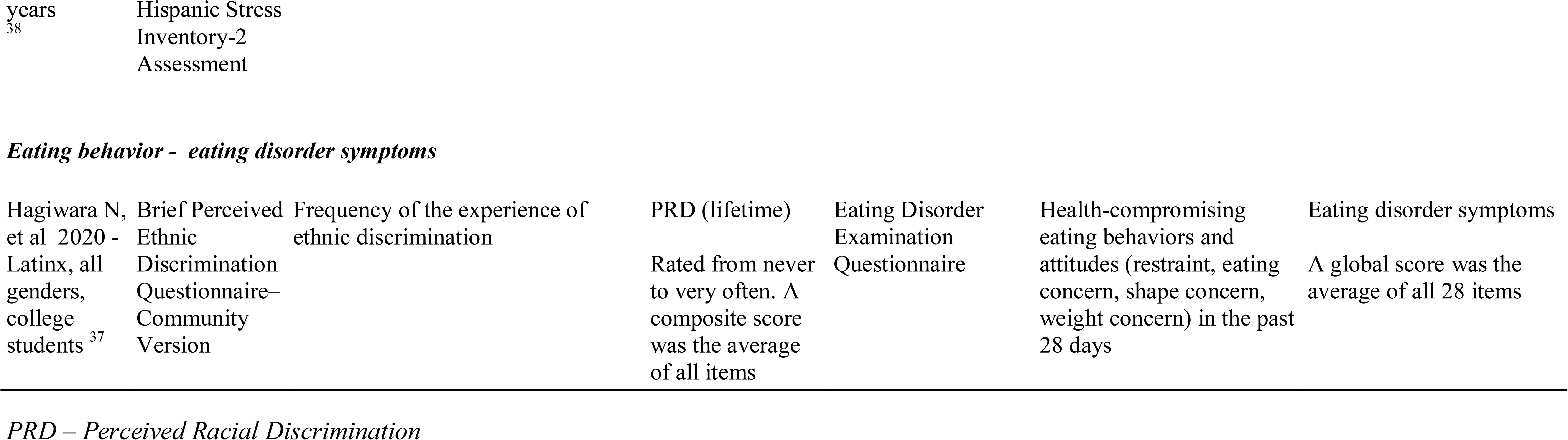
Characteristics of exposure and outcome measurement according to type of outcome (n-19).

| **Exposure (perceived racial discrimination)** |  |  | **Outcome (eating habits)** |  |  |  |
| --- | --- | --- | --- | --- | --- | --- |
| ID - setting | Instrument | Assessment | Exposure | Instrument | Assessment | Outcome |
| *Food consumption – nutrient/food group consumption* |  |  |  |  |  |  |
| Corral I. & Landrine H., 2012 - Black/African-American, 18-95 years<sup>28</sup> | Non validated questionnaire | Amount of PRD experienced recently | PRD (past year)<br>Dichotomous: low or high | Non validated 5-a-day fruit and vegetable survey | Weekly consumption of juice, fruit, salad, and beans | Fruit and vegetable intake<br>Dichotomous: $\geq 5$ or $< 5$ servings of fruits and vegetables a day). |
| Cuevas A., et al 2014 - Black/African-American, 18 years or more<sup>13</sup> | Day-to-Day Unfair Treatment Scale | Frequency of discrimination experienced daily in different settings based on race/ethnicity or skin color. | PRD (lifetime)<br>Higher scores were indicative of greater PRD | NCI 5-A-Day fruit and vegetable questionnaire | Fruits and vegetables consumption | Fruit and vegetable intake<br>Dichotomous: $\geq 5$ or $< 5$ servings of fruits and vegetables a day). |
| Sims M, et al 2016 - Black/African-American, 35-84 years<sup>32</sup> | JHS Discrimination Instrument | Frequency of racial discrimination experienced daily (everyday discrimination) and occurrence of unfair treatment over the lifetime (lifetime discrimination) in different settings | PRD (lifetime)<br>Scored such that higher values indicate more everyday and lifetime PRD | NA (not available) | Percent daily calories from fat - men | Fat consumption<br>NA |
| Nadimpalli S, et al 2017 | Everyday Discrimination | Frequency of interpersonal discrimination experienced, unfair | PRD (past year) | Food Frequency Questionnaire (FFQ) | Frequency of fruit/vegetable, American | Fruit/vegetable and sweets intake |
| - South Asians, 40-79 years<sup>34</sup> | Scale | treatment, and discrimination-related hassles. | The scale ranges from 9 to 54 with higher values indicating more discrimination. |  | and south Asian sweets consumption over the past year | Higher scores indicating higher frequencies of fruit/vegetable, American and South Asian sweets consumption |
| ***Food consumption – diet quality*** |  |  |  |  |  |  |
| Brodish A, et al 2011 - Black/African-American, 18-95 years<sup>24</sup> | Non validated questionnaire | Amount of PRD at work or in college (age 19 and 21) | PRD (lifetime)<br>Higher scores were indicative of greater PRD | Non validated diet quality survey | Frequency of vegetables, fast food, sweets and snacks consumed at age 30. | Healthy diet<br>Higher values indicate a more healthy diet. |
| Forsyth J, et al 2014 - Black/African-American, hypertension, 18 years or more<sup>25</sup> | The Schedule of Racist Events (SRE) | Frequency of discrimination experienced in different settings in the past year; over the lifetime; and the appraisal of the experiences as stressful | PRD (lifetime)<br>PRD (past year)<br>PRD (stress)<br>Scored from 18 to 108. Higher scores indicating more frequent exposure to PRD | Rapid Eating and Activity Assessment for Patients (REAP) | Baseline and study time intake of fruits and vegetables, whole grains, sodium, fat, saturated fat, cholesterol, calcium-rich foods, and sugary foods and drinks. | Unhealthy diet<br>Three-point Likert format (1=often to 3=rarely/never) and higher scores indicating less healthy eating habits. |
| Dawson A, et al 2015 - Black and White people, adults, diabetes, 18 years or | DISTANCE survey | Frequency people were treated poorly or made feel inferior because of race/ethnicity within the past year. | PRD (past year)<br>Rated from never to often | Diabetes Self-Care Activities (SDSCA) scale | Frequency of self-care activity in the last 7 days for general diet (follow healthy diet) and specific diet (i.e. fruits and fats) | Healthy diet<br>Not informed |
| Simons R, et al 2020 - African American, men and women , 40-60 years<sup>26</sup> | The Schedule of Racist Events (SRE) | Frequency of discrimination experienced in different settings in the past year | PRD (past year)<br>Higher scores were indicative of a higher frequency of PRD | Non validated healthy diet scale | Frequency of fruits, vegetables, fatty and fast foods consumption in the past seven days | Healthy diet<br>Higher scores indicating a more healthy diet |
| Kohlmaier J. 2004 - Black/African-American, 18 - 60 years<sup>30</sup> | The Schedule of Racist Events (SRE) | The degree to which racist discrimination is evaluated or appraised as stressful | PRD (stress)<br>Higher scores are indicative of more experience with racism. | Binge Eating Scale (BES) | Behavioral aspects of binge eating | Binge eating<br>Higher scores indicate more binge eating |
| Harrington E, et al 2006 - Black and White people, women, undergraduate students, 21 years (mean age)<sup>35</sup> | The Schedule of Racist Events (SRE) | Frequency of discrimination experienced in different settings in the past year; over the lifetime; and the appraisal of the experiences as stressful | PRD (stress)<br>Higher scores were indicative of greater PRD | Binge Eating Scale (BES) | Binge Eating Symptomatology | Binge eating<br>Scores range from 0 to 46, with higher scores indicating greater severity of binge eating symptomatology |
| Clark J & | The Schedule | Frequency of discrimination | PRD (lifetime) | Binge Eating Scale | Presence of binge eating | Binge eating |
| Winterowd C, 2012 - Native Americans, 19 - 70 years<sup>33</sup> | of Racist Events (SRE) | experienced in different settings in the past year; over the lifetime; and the appraisal of the experiences as stressful | PRD (stress)<br>Higher scores were indicative of greater PRD |  | behaviors, feelings and cognitions associated with binge eating | Rated from 0 (no binge eating problem) to 3 (severe binge eating problems). |
| Johnson P, et al 2012 - Black/African-American, women, weight control, 18-71 years<sup>14</sup> | The Perceived Discrimination Questionnaire | Frequency of discrimination experienced in different settings based on race/ethnicity or skin color. | PRD (lifetime)<br>Higher scores were indicative of greater PRD | Non validated emotional eating survey | Frequency of eating motivated by depression or sadness and frequency of eating motivated to manage stress | Emotional eating<br>Rated from (1) never to (7) refused to answer question |
| Hoggard L, et al 2019 - Black/African-American, undergraduate students, 18 - 27 years<sup>29</sup> | Racism and Life Experience scale (The Daily Life Experiences subscale) | The frequency with which the participants experienced 18 racial hassles in the past year | PRD (past year)<br>Higher scores were indicative of greater PRD | The Emotional Eating Scale (EES) - Angry, Worried and Sad subscales | The urge to cope with negative affect by eating | Emotional eating<br>Higher scores are indicating a greater urge to cope with negative affect through eating. |
| Longmire-Avital B & McQueen C, 2019 - Black/African | Index of Race Related Stress, adapted | The number of chronic stress experienced as a result of racist acts | PRD (stress)<br>Not informed | Eating Behavior Patterns Questionnaire (eating habits subscale) | Eating habits | Emotional eating<br>Rated on a Likert scale from 1 (strongly disagree) to 5 (strongly agree). |
| Connolly M, 2011 - Black/African-American, 18-73 years<br>27 | Everyday Discrimination Scale | Frequency of racist events experienced in the past year | PRD (past year)<br>Higher scores were indicative of greater PRD | Yale Eating Pattern Questionnaire (YEPQ) | Inhibition around food, which may lead to overeating (Uninhibited scale)<br>Restraint around food and overeating (Oversnacking and Binging scale)<br>Inhibition and restraint around food, leading to overeating (Total scale) | Patterns of overeating<br>Higher scores indicate lower levels of inhibition and restraint around food, which may lead to overeating. Total scores are obtained by reversing scores on two items within the Uninhibited scale and then summing scores across all scale items. |
| Hayman L, 2012 - Black/African-American, women, undergraduate students, 18 years or | The General Ethnic Discrimination Scale | Frequency and the appraisal of reports of discriminatory events. | PRD (past year)<br>PRD (lifetime)<br>Higher scores were indicative of greater PRD | Dutch Eating Behavior Questionnaire (restrained and emotional eating subscales) | Frequency of dieting behaviors enacted in efforts to either maintain or lose weight (Restraint eating subscale)<br>Frequency of desire to eat while experiencing specific emotions, such as when | Restrained and emotional eating<br>High scores on the restraint measure indicate that participants often try to restrain their food intake. High scores on the |
| Kelly N, et al 2018 - African-American, asian and hispanic, men, 18-30 years <sup>36</sup> | Everyday Discrimination Scale Asian American Racism-Related Stress Inventory<br>Discrimination Stress subscale of the Hispanic Stress Inventory<br>Non validated questionnaire to assess post-election PRD | Frequency of discrimination experienced in different settings based on race/ethnicity or skin color, usually and postpresidential elections | PRD (past year)<br>PRD (post-election)<br>Higher scores were indicative of greater PRD | Eating Disorder Examination Questionnaire | Frequency of loss of control eating | Loss of control eating<br>Not informed |
| Kelly N, et al 2020 - African-American, asian and hispanic, men, 18-30 | Everyday Discrimination Scale, Asian American Racism-Related Stress Inventory, | Frequency of discrimination experienced in different settings based on race/ethnicity or skin color | PRD (past year)<br>Higher scores were indicative of greater PRD | Eating Disorder Examination Questionnaire | Frequency of loss of control eating | Loss of control eating<br>Not informed |
| Hagiwara N, et al 2020 -<br>Latinx, all<br>genders,<br>college<br>students <sup>37</sup> | Brief Perceived<br>Ethnic<br>Discrimination<br>Questionnaire–<br>Community<br>Version | Frequency of the experience of<br>ethnic discrimination | PRD (lifetime)<br>Rated from never<br>to very often. A<br>composite score<br>was the average<br>of all items | Eating Disorder<br>Examination<br>Questionnaire | Health-compromising<br>eating behaviors and<br>attitudes (restraint, eating<br>concern, shape concern,<br>weight concern) in the past<br>28 days | Eating disorder symptoms<br>A global score was the<br>average of all 28 items |

*PRD – Perceived Racial Discrimination*

### Eating habits

Eight studies investigated food consumption as the outcome ^8, 13, 24–26, 28, 32, 34^. Four studies observed the intake of specific food groups or nutrients as a proxy of healthy or unhealthy food consumption ^13, 28, 32, 34^ like fruit and vegetables ^13, 28, 34^, fats ^32^, and sweets ^34^. In these studies, the frequency of consumption of unhealthy or healthy foods indicated the quality of diet. Moreover, four studies focused on diet quality ^8, 24–26^, applying a food frequency questionnaire (FFQ) for specific food items, like vegetables, fruits, whole grains, fast foods, sweets, and snacks. From these instruments, they created healthy or unhealthy eating indexes. The authors used validated ^8, 13, 25, 34^ and non-validated ^24, 26, 28^ instruments to measure food consumption (Table 3).

The majority of the studies (n=10) investigated eating behaviors ^4, 14, 27, 29–31, 33, 35–37, 38^. Emotional eating was assessed in 4 studies ^4, 14, 29, 31^, and binge eating appeared in 3 studies ^30, 33, 35^. Other eating behavior outcomes, like overeating, restraint, eating, and body concern were assessed less frequently. Emotional eating was measured by validated instruments ^4, 29, 31^, and one non validated emotional eating survey ^14^. While some researchers investigated the desire to eat while experiencing specific emotions as a whole, some studies evaluated and sorted particular emotions to understand the motivations to eat. Some examples are emotional distress ^30^, depression ^14, 29^, anxiety or anger ^29^, sadness ^14^, and the frequency of eating to manage stress ^14^. Loss of control eating appeared in two studies ^36, 38^.

### PRD and eating habits

Table 4 presents the main results of the association between PRD and eating habits. Of the 19 studies included in this review, 46 associations were established. Of these, 38 associations revealed that PRD is associated with negative eating habits. Seven associations were null ^13, 27, 28, 34, 35, 38^. Only 1 study found that PRD is associated with positive eating habits ^25^.

**Table 4.**
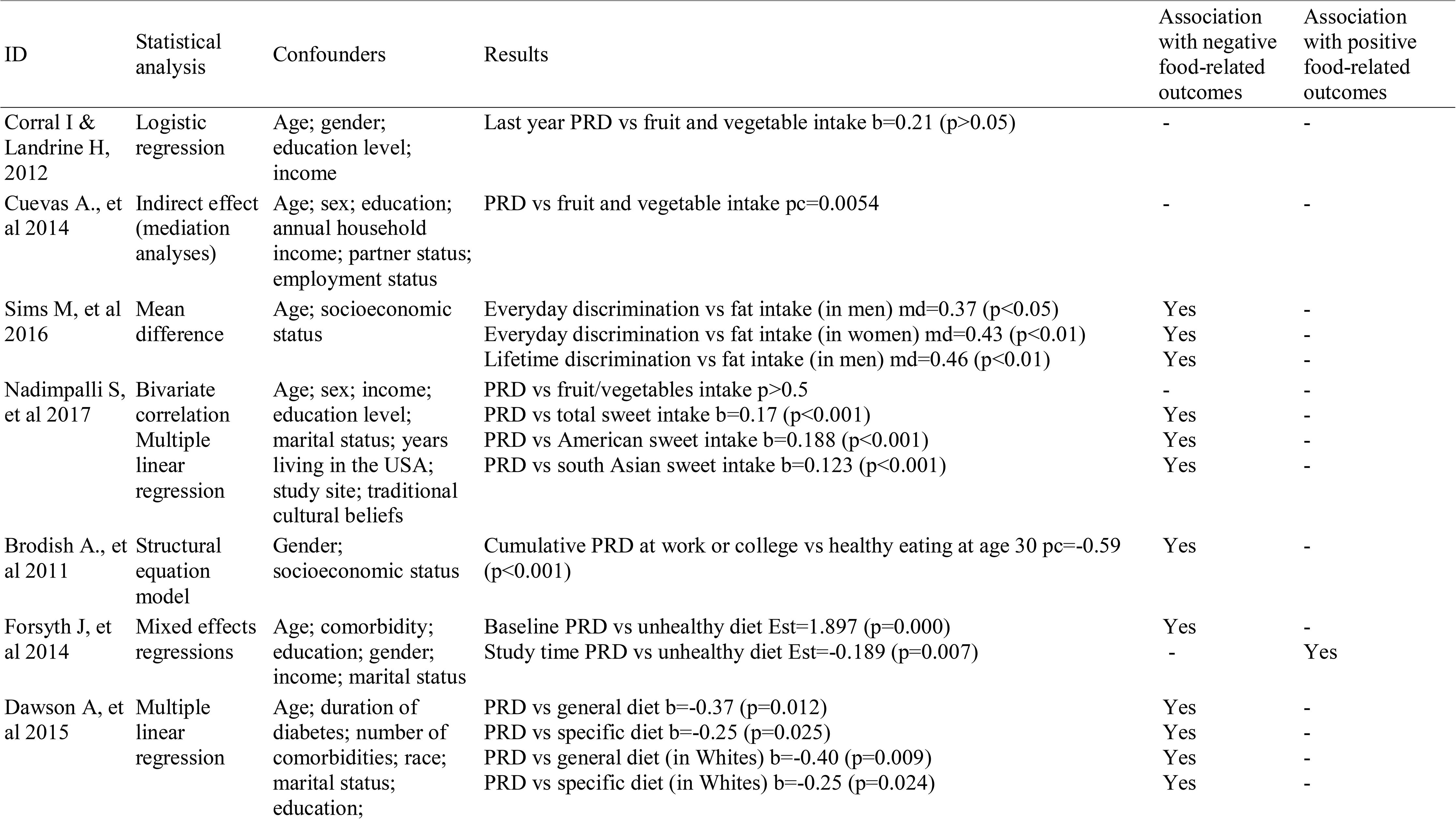
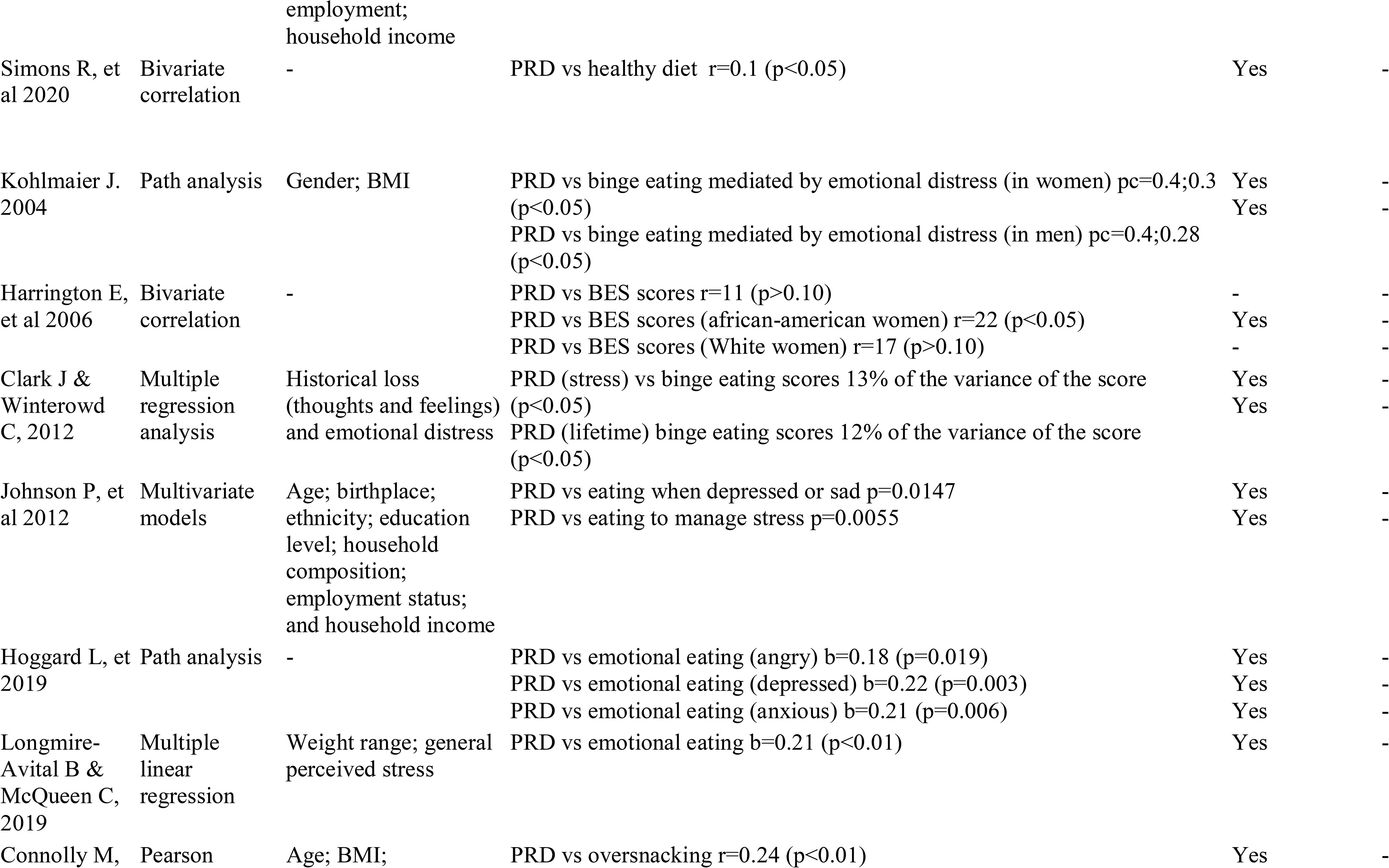
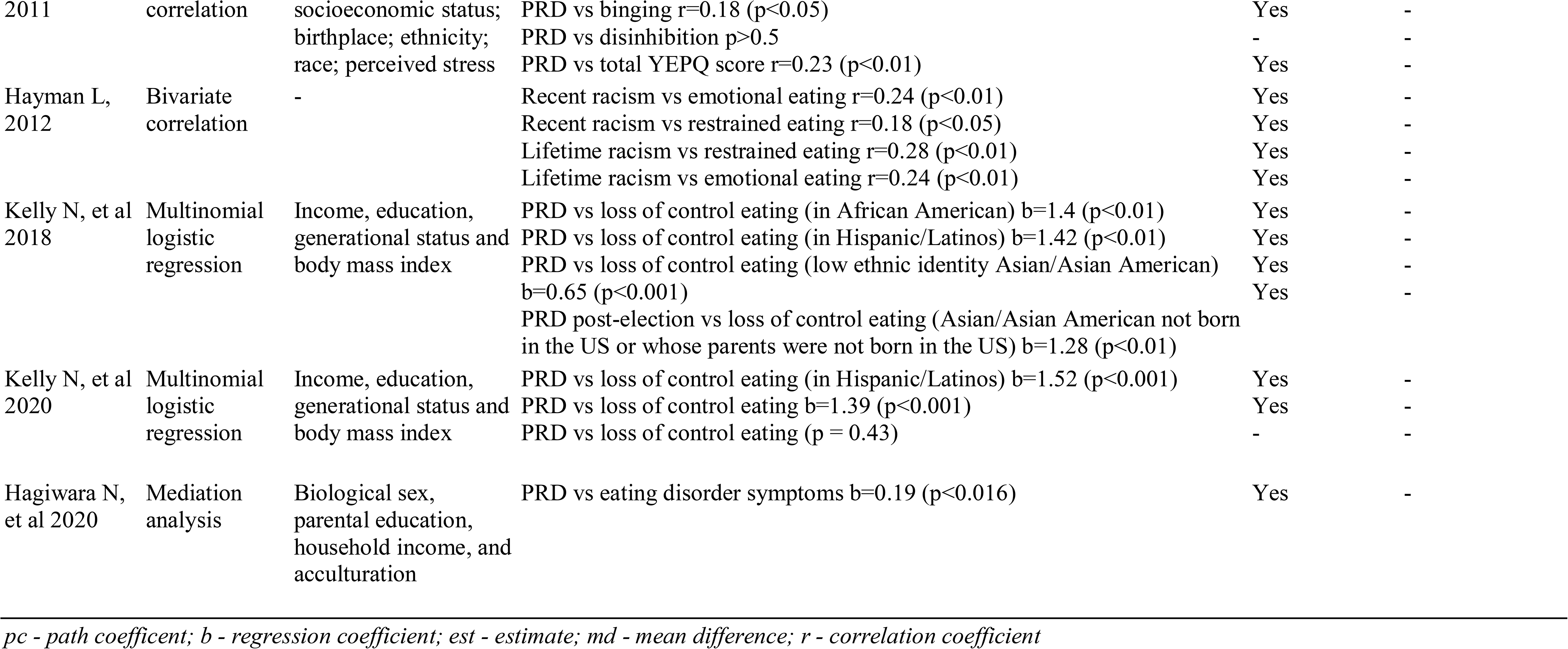
Main results of the association between perceived racial discrimination and eating habits (n=19)

Summing up the main findings of PRD and food consumption outcomes, there were 17 associations in total. Thirteen of them show a relationship between PRD and unhealthy eating ^8, 24–26, 32, 34^, 3 of them were null ^13, 28, 34^, and only one is related to a healthy diet ^25^. One study included both Black/African-American and White people, demonstrating that a worse diet among Whites is associated with PRD ^8^. All cohort studies belonged to the food consumption outcome group. One of them presents divergent data: a change in diet from baseline to 12 months that was significantly greater for those with high PRD exposure than those with low PRD. It means that greater exposure was associated with more remarkable improvement in the diet throughout the study ^25^.

Regarding eating behavior outcomes, 29 associations were established. There were 8 positive associations with emotional eating ^4, 14, 29, 31^, 6 with binge eating ^27, 30, 33, 35^, 6 with loss of control eating ^36, 38^, two with restrained eating ^4^, two with over-snacking and disinhibition ^27^, 1 with health-compromising eating behaviors and attitudes (restraint, eating concern, shape concern, weight concern) ^37^. Four associations were null ^13, 28, 34, 38^. One study included both Black/African-American and White women, demonstrating that binge eating among Black women is affected by PRD. In contrast, the Binge Eating Scale (BES) scores among Whites did not seem to be associated with PRD. Four studies include both genders, and the results were similar for both ^35^.

Demographic (age, sex, gender, birthplace, study site, live in the USA, generation), socioeconomic (race/ethnicity, income, employment status, education level, parental education, marital status, household composition), psychological (thoughts, emotions, stress, beliefs), related to nutritional status (BMI and weight change) and acculturation variables were included in analyses as confounders or mediators. The majority of the studies performed a multivariate analysis (42%). Five studies performed path analysis ^13, 24, 29, 30, 37^.

### Conceptual causal diagrams of associations between PRD and food habits

The primary studies formulate two main conceptual models of the association between racial discrimination and health outcomes, as shown in Figure 2. The first model establishes that chronic stress due to everyday perceived racial discrimination predicts alterations in food consumption and eating behavior. These alterations in food habits, in the long run, clearly lead to poor health outcomes ^8, 25, 36^. The second model includes the previous pathway and suggests that stress can directly affect health outcomes (the dashed line represents it in Figure 2) _26,29,31_.

**Figure 2.**
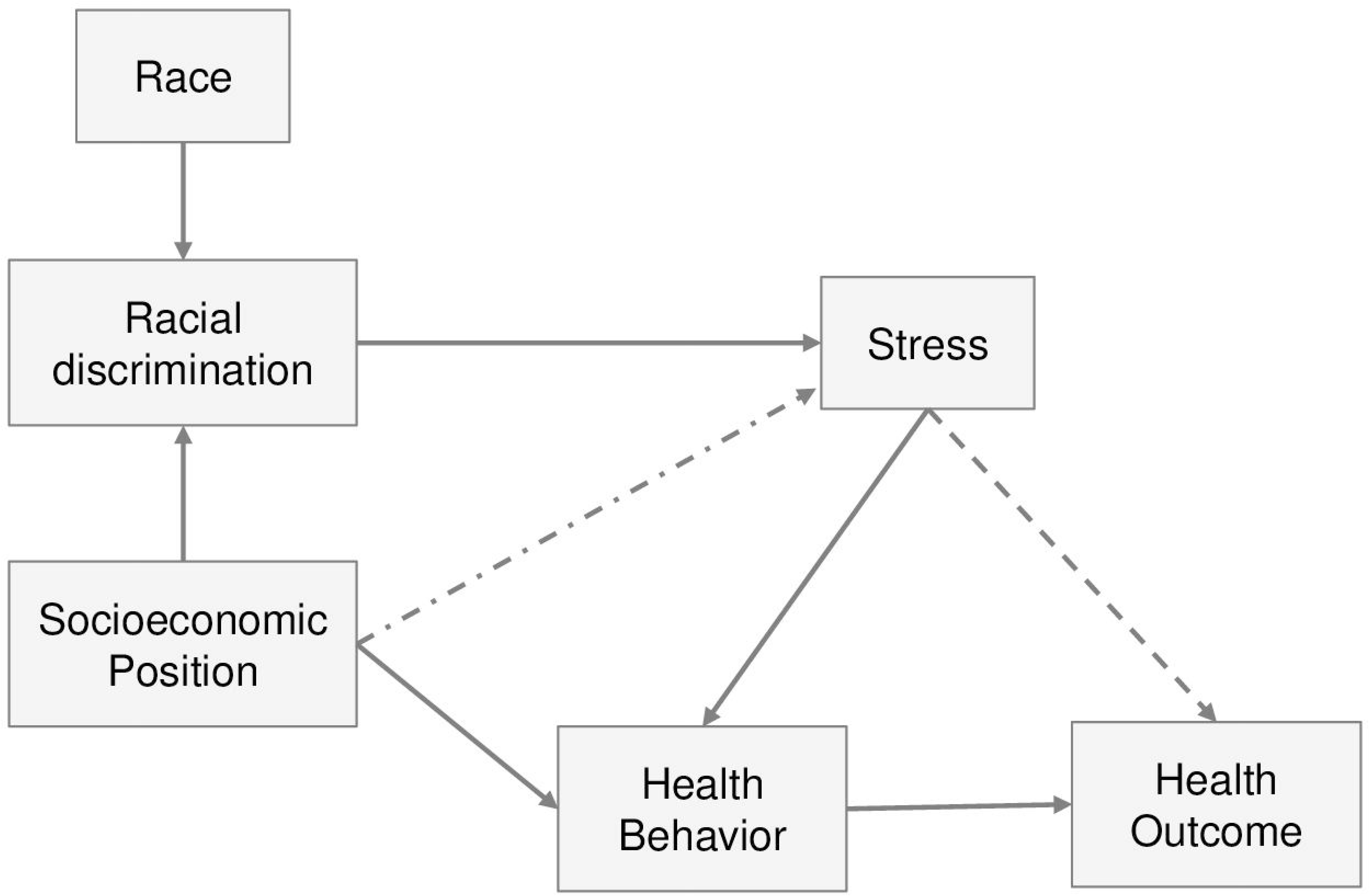
Main conceptual models of the association between Perceived Racial Discrimination (PRD) and health outcomes

Two additional observations must be made to the model. All the authors are prone to claim that race and ethnicity are highly correlated with different group experiences in societies; racial discrimination has been one of these experiences. Also, many studies suggest that racial discrimination experience intersects with the socioeconomic position of individuals (gender, age, education, income, ethnic identity) and other cultural and historical factors (racial socialization, social and parental support, cultural constructs and stereotypes, social norms, and historical traumas) ^24, 33, 36, 37^.

## Discussion

### Summary of evidences

The present study aimed to examine the available evidence about the relationship between PRD experiences and eating habits. The majority of the studies included showed that PRD was associated with poor food habits, including unhealthy food consumption (consumption of sweets and fats and lower consumption of fruit and vegetables), emotional eating, binge, loss of control, dietary restraint, over-snacking, eating disorders symptoms, and disinhibition.

### Providing possible explanations for the association between PRD and food habits

Racism and racial discrimination can be defined and measured at different levels, such as internalized, interpersonal, institutional, and structural. Internalized discrimination can be defined as theincorporation and acceptance, consciously or unconsciously, of a racist and inferior image of oneself from a dominant and privileged group. Interpersonal racial discrimination is defined as the experience of receiving unfair treatment, usually with expressions of beliefs, stereotypes, and prejudices, in interpersonal encounters. Institutional racism is the manifestation of racial discrimination through formal systems, usually legal and social institutions, such as criminal justice, the distribution and disparities of educational and health opportunities or labor and housing markets, or more explicit discriminatory policies ^1, 5^. Finally, structural racism is a much more comprehensive framework that encompasses different dimensions of racism, from the interpersonal to the cultural and formal, state-levels of racial discrimination. It is sustained by a racially-hierarchical culture of values, beliefs, social practices, norms and interconnected inequitable systems (such as legal, cultural, and social institutions) that allow and/or encourage systematic oppression and unequal treatment to specific groups ^5, 39, 40^.

Although most studies assumed the need to study all these levels of racism is essential to provide insights into the association of racism and health outcomes, all of them focused on questions and measurement instruments for interpersonal racial discrimination. Nonetheless, few studies included structural and cultural constructs in their analysis with broader conceptual models to explain their results ^24, 29, 30, 33, 37^.

Perceived racial discrimination is the central scientific construct to measure racial discrimination. Therefore, the chronic stress due to racial discrimination is based only on perceived discriminations in interpersonal encounters, during the individuals’ lifetime, or in a specific time frame or setting. Self-reports of perceived discrimination are the most common discrimination measures used in psychology and epidemiological fields. They aim to capture an individual’s appraisal of discriminatory treatment. This appraisal process determines whether the treatment is evaluated as stressful or as a threat that taxes or exceeds an individual’s available coping resources. Thus, perceived discrimination is considered a biopsychosocial stressor for the targeted individual ^41^.

In order to explain the association between perceived racial discrimination and health and behavior outcomes, all studies included in this review share one main argument: chronic stress affects health behaviors and health outcomes. Everyday exposure to racial discrimination thought life is a specific cause of chronic stress. Thus, racial discrimination affects health behaviors and health outcomes. To fully understand the presuppositions and implications of this conclusion, we need to unpack its central concepts and the underlying mechanisms.

Discriminatory events can trigger interplayed stress responses, usually centered on physical, psychological, social, and cultural issues. The physiological mechanism is explained through the neuroendocrine stress response. Chronic stress leads to activation of the hypothalamus-pituitary-adrenal axis that affects both health outcomes and health behaviors. In one way, activation of the hypothalamus-pituitary-adrenal axis causes an inflammatory process mediated by glucocorticoid hormones (cortisol and corticosterone). Abnormal cortisol secretion can promote obesity in two ways. First, excess cortisol concentrations have been associated with visceral fat accumulation. One explanation for this may be increased glucocorticoid metabolism due to increased glucocorticoid receptor density in intra-abdominal adipose tissue compared to other regions. Glucocorticoids also affect visceral fat via their effect on lipid metabolism. Acutely, physiological cortisol concentrations stimulate whole-body lipolysis. In the presence of insulin, increased cortisol concentrations inhibit lipid mobilization and favor lipid accumulation either directly by stimulation of lipoprotein lipase or indirectly by inhibiting the lipolytic effects of growth hormone ^42^. This can affect weight gain without necessarily changing food consumption/or eating behavior. Second, chronic activation of the HPA axis alters glucose metabolism, promotes insulin resistance, with changes in several appetites related hormones (e.g. leptin, ghrelin) and feeding neuropeptides (e.g. NPY). This causes changes in the mechanisms of hunger and satiety. Thus, elevated cortisol secretion can lead to chronically stimulated eating behavior, enhancing the propensity to eat high-calorie palatable food. This relationship is modulated by a reward system, as well. Reward circuitry could interact with the HPA axis, meaning eating high-palatable foods leads to a partial suppression of the HPA axis and cortisol secretion. A bidirectional relationship between cortisol levels and behavioral eating would constitute a feedback loop when these foods’ consumption signals a decrease in stress upon reaching satiety ^43^.

In this sense, despite none of the studies included in this review had evaluated cortisol levels, several studies in adults have been described the association between perceived discrimination, including PRD, and a flatter diurnal cortisol slope, i.e., less sleep decline throughout the day, with lower morning levels and higher evening levels. ^44, 45^ A prospective study reported that greater PRD measured over 20 years predicts flatter diurnal cortisol slopes later in life ^46^.

Regarding the psychological mechanisms addressed by the articles, the authors mention the stress and coping framework, especially. Individuals may appeal to coping strategies to alleviate stress. The psychological response to stress due to discrimination events can result in different adverse emotions, such as anxiety, depression, guilt, anger, sadness, loneliness, low self-esteem, and helplessness ^47^. Coping strategies can be adaptive when the individual uses healthy ways to deal with these experiences, such as searching for family and community support, rest, and physical activity. Maladaptive coping occurs when, to manage stress and negative emotions, the discriminated person engages in negative health behaviors or decreases the ability to promote health promotion behaviors. The resulting changes in health behaviors are ultimately related to poor health outcomes. It is important to highlight that in the latter case, the feeling of relief is transitory and may be accompanied by emotions such as guilt and frustration afterward. Maladaptive coping, thus, may improve mental health in the short term, but it can be problematic and harmful in a long time to psychological and physical health ^48^. It is important to underline that these behavioral-coping strategies can be influenced by differences in the person’s social role and the cultural and social norms embedded in it ^36^. In Connolly’ study, positive racial socialization protects young Black women from developing an eating disorder by correcting a false, incomplete and misunderstood interpretation of what it means to be Black in the United States. Positive racial socialization and feelings of belongingness alleviates the harms of interpersonal and cultural racial discrimination and/or lead to adaptive health behaviors, such as physical activities and healthy diets ^27^. On the other hand, racial socialization and/or ethnic self-identification do not protect from maladaptive behaviors in all cases. Clark et al. study showed that ethnic self-identification and acculturation of Native American/American Indian women might be a predictor for binge-eating behaviors due to feelings of historical trauma or historical loss that has been passed from generations ^33^.

There are individual and collective factors that can contribute to the different impacts of racial discrimination on health behaviors among individuals. The Theory of Cognitive Appraisal postulates that the negative consequences of discrimination depend firstly on whether the stressor is relevant and dangerous and secondly on whether the victim does not have sufficient resources to contain the stressor. The perception of social support is a resource that can prevent or hinder maladaptive coping ^49^; as well as emotional self-regulation and cognitive resources ^50^.

On a broader level, social class may influence the intensity of exposure and the kind of interpersonal racial discrimination people of color face during their lifetime and, consequently, their health outcomes. Social class can be understood as a relational concept, expressing the interdependent economic and legal relationship between groups of people ^51^. This structural location of the individuals within the economy may influence the distributions of resource-based goods (such as income, wealth, education, and neighborhood) and prestige-based good (such as social status, access to goods, services, knowledge, prestige, income, and educational level). Several studies explicitly acknowledge that class is a social determinant for their results, but it is restricted to discussing their limitations ^8, 13, 24, 29, 34^. The socioeconomic context of individuals, for example, plays an important role in structural and cultural dimensions, such as nutrition policies, food environment, and food supply, and health care access, and acculturation of food practices (43,44).

In this way, it is possible to conclude that weight gain and development of NCD do not depend exclusively on individual behaviors or food choices. The degradation of biological systems through the aforementioned physiological mechanisms is called the allostatic charge ^52^. As in this model, epigenetic indexes have been developed to assess discrimination’s cumulative effect, considering all discrimination levels ^53^. In this way, assigning the responsibility to adopt healthy eating behaviors in the victims of racial discrimination, overestimating the agency of these population groups to promote this type of change. The food environment is a fundamental component to understand how racial discrimination affects individuals’ consumption and eating behavior. Food deserts are characterized by neighborhoods that do not have establishments that sell fresh or minimally processed food in sufficient quantity or adequate quality. Food deserts are usually located in poor neighbourhoods, where the population with the lowest socioeconomic status is concentrated^54^

Not only the food environment but the neighborhood as a whole can favor or prevent the adoption of healthy behaviors, such as safety, paved streets, trees that provide shade. Even when the food environment consists of establishments that sell quality food, financial access to these products is a barrier to consuming fresh or minimally processed foods. The same occurs in neighborhoods where there are gyms and clubs for physical activity, which are usually expensive and inaccessible to a large part of the population. The emphasis on a healthy lifestyle ignores the importance of adverse health determinants. These social conditions can only be addressed through social policies and programs that target institutional racism and promote economic equality ^55^.

Some studies have found no association between self-perceived racial discrimination and eating habits ^13, 28, 34^. Cuevas (2014) examined whether stress or depressive symptoms mediated the association between self-perceived discrimination and multiple behavioral risk factors for cancer among blacks, such as inadequate diet. There was also no association with the consumption of fruits and vegetables ^13^. Nadimpalli (2017), in a study conducted with South Asians, also found no association with the consumption of fruits and vegetables ^34^.

One possible explanation given is that this can be attributed to the fact that groups discriminated by race/ethnicity do not necessarily eat less healthy food, but eat more unhealthy food. Another explanation is that interpersonal racial discrimination may be responsible for greater consumption of comfort food to deal with stressful experiences while structural discrimination is more related to the low consumption of fresh and minimally processed foods, due to barriers of physical access, lack of sufficient and high quality food.

### Proposing further elements to develop an integrated conceptual model

Most of the studies claim that investigating different levels of racial discrimination should provide new insights into the association of discrimination and health outcomes. However, taking all these levels into account can be hard to unite into a theoretical and explanatory framework, not to mention their measurement. Based on the ecosocial theory formulated by Nancy Krieger (2011), we propose a new and more complex conceptual model (Figure 3) to guide future researches. Whereas the studies under review are based fundamentally on biopsychosocial theories, the ecosocial theory allows us to incorporate different, broader levels of phenomena in a single framework. It takes the multilevel notion of embodiment as a central scientific construct to understand social, material, and ecological contexts that we live. Thus, as we can see in the model, the main focus is to discriminate the multiple pathways that racial discriminations are biologically incorporated in individuals’ bodies and affect the patterns of health outcomes and health behaviors.

**Figure 3.**
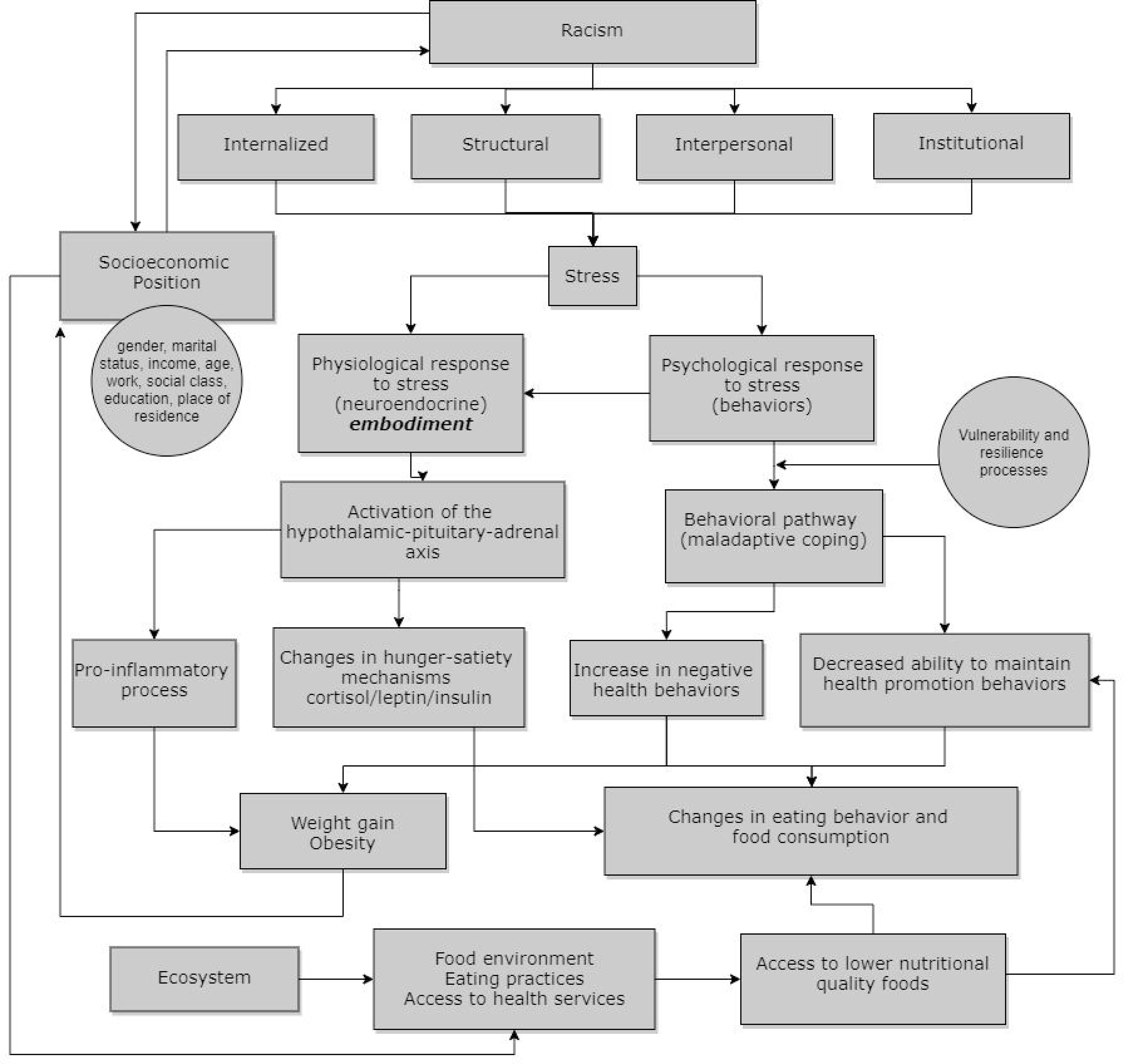
Conceptual model based on the association between Perceived Racial Discrimination (PRD) and eating habits

Along with interpersonal discrimination, we can have institutional and structural racism. The model synthesizes the discussions made above. The main deficit in the studies under review is the assessment and measurement of historical and contextual dimensions, which should be better investigated to establish how specific determinants can influence different groups.

### Strengths and limitations

This study represents an innovative and socially relevant addition to the field of food and nutrition. It was conducted according to the main guidelines for systematic reviews and, to the best of the author’s knowledge, this is the first one on the topic. In addition, a new conceptual model is formulated to integrate the available social epidemiology and psychology theories to explain the mechanisms of association.

As pointed out in previous publications, there are methodological limitations inherent in studies that assess food habits. Regarding food consumption, some studies used methods such as FFQ or food diaries and may be more accurate than others, such as simple questions about food group consumption. Also, different methods were used for data collection of eating behavior. Concerning the exposure, most studies used validated instruments to measure perceived racial discrimination, resulting in an accurate measurement of it. In addition, there is a risk of bias inherent in the observational design of the studies because interventions are not feasible given the nature of the exposure, which cannot be observed isolated from its social context. Finally, while longitudinal studies are well known for being a better study design to investigate the association’s direction, most of the studies selected for this review had a cross-sectional design, limiting the temporality evidence of the results presented in this review.

Finally, it is important to highlight that discrimination based on race and ethnicity has complex cultural, historical and intersectional aspects. The experience of discrimination is lived in different ways for different racial groups. It must be considered that racism experienced by African Americans is different from the racism or ethnic discriminations that occurs with Asians, Native Americans, or Latinos living in the United States, for example. Thus, these aspects must be considered when studying racism.

## Conclusions

Racial discrimination appears to be associated with unhealthy eating habits. The articles retrieved in this systematic review revealed that victims of enduring, day-to-day racial discriminations exhibit poor eating behavior and food consumption. This can be represented by the consumption of sweets and fats and lower consumption of fruits and vegetables, emotional eating, binge eating, loss of control when eating, food restriction, overeating, disinhibition and other symptoms of eating disorders. Everyday exposure to racial discrimination through life is a specific cause of chronic stress that is biologically and psychologically incorporated into our bodies. The biopsychosocial framework was the dominant theoretical approach used to formulate the main hypothesis and explain the causal pathways of the studies.

Nonethless, only a biopsychosocial framework to understand the perils of racism is not enough. It was proposed a new conceptual model based on ecosocial framework to guide future researches. Frameworks and conceptual models are important tools in scientific research, because they are a guide to formulate questions and hypothesis, construct concepts and explane causal pathways of disease distribution. The domain of the ecosocial framework allows us to integrate different domains of exposures and the interactions between the social and the biological processes. Thus, different levels of determination of food habits should be taken into account to understand the effects of health inequities and build more efficient public policies. Finally, the research agenda on this issue needs more longitudinal studies in order to confirm the association demonstrated by this review. Also, studies in different countries (beyond USA) should be conducted to understand how racial discrimination affects people’s food behavior of different ethnic, cultural, social, and economic contexts.

## Data Availability

none

## Notes

### Competing Interest Statement

The authors have declared no competing interest.

### Author Declarations

This is a systematic review

