## Supplemental Tables for "Perceived racial discrimination and eating habits: Systematic review and conceptual models"

Supplemental Table 1. Characteristics of the studies (n=19).

| ID | Study design | Data collection timeframe | Population | Sample size | Age |
|---|---|---|---|---|---|
| ***Food consumption*** | | | | | |
| Brodish A., et al 2011 | Prospective cohort | 1991-2009 | Black/African-American | 815 | 12-30 years |
| Corral I & Landrine H, 2012 | Cross-sectional | Not available | Black/African-American | 2190 | 18-95 years |
| Cuevas A., et al 2014 | Prospective cohort | 2008-2009 | Black/African-American | 1365 | 18 years or more |
| Forsyth J, et al 2014 | Prospective cohort | Not available | Black/African-American, hypertension | 930 | 18 years or more |
| Dawson A, et al 2015 | Cross-sectional | 2014 | Black and White people, adults, diabetes | 602 | 18 years or more |
| Sims M, et al 2016 | Cross-sectional | 2000-2004 | Black/African-American | 4925 | 35-84 years |
| Nadimpalli S, et al 2017 | Cross-sectional | 2010-2013 | South Asians | 906 | 40-79 years |
| Simons R, et al 2020 | Prospective cohort | 2005-2008 | Black/African-American | 494 | 40-60 years |
| ***Eating behavior*** | | | | | |
| Kohlmaier J. 2004 | Cross-sectional | Not available | Black/African-American | 385 | 18 - 60 years |
| Harrington E, et al 2006 | Cross-sectional | Not available | Black and White people, women, undergraduate students | 178 | 21 years (mean age) |
| Connolly M, 2011 | Cross-sectional | 2009 | Black/African-American | 201 | 18-73 years |
| Clark J & Winterowd C, 2012 | Cross-sectional | 2007 | Native Americans | 269 | 19 - 70 years |
| Hayman L, 2012 | Cross-sectional | 2010 | Black/African-American, women, undergraduate students | 319 | 18 years or more |

| | | | | | |
|---|---|---|---|---|---|
| Johnson P, et al 2012 | Cross-sectional | 2003 | Black/African-American, women, weight control | 350 | 18-71 years |
| Kelly N, et al 2018 | Cross-sectional | 2017 | African-American, asian and hispanic, men | 780 | 18-30 years |
| Hoggard L., et al 2019 | Cross-sectional | 2014-2017 | Black/African-American, undergraduate students | 150 | 18 - 27 years |
| Longmire-Avital B & McQueen C, 2019 | Cross-sectional | 2014 | Black/African-American, women, undergraduate students | 149 | 18-25 years |
| Kelly N, et al 2020 | Cross-sectional | 2017 | African-American, asian and hispanic, men | 798 | 18-30 years |
| Hagiwara N, et al 2020 | Cross-sectional | - | Latinx | 198 | 18-25 years |

Supplemental table 2 - Methodological quality assessment (n=19).

| Studies | Selection | | | | Comparability | Outcome/exposure | | | Total | % |
|---|---|---|---|---|---|---|---|---|---|---|
| | 1 | 2 | 3 | 4 | 5 | 1 | 2 | 3 | | |
| ***Food consumption*** | | | | | | | | | | |
| Brodish A., et al 2011† | ☆ | ☆ | ☆ | ☆ | ☆ | - | ☆ | ☆ | 7 | 87,5 |
| Corral I & Landrine H, 2012 | ☆ | ☆ | ☆ | ☆ | ☆☆ | ☆ | ☆ | - | 8 | 80 |
| Cuevas A., et al 2014† | - | ☆ | ☆ | - | ☆ | - | - | - | 3 | 37,5 |
| Forsyth J, et al 2014† | ☆ | - | - | ☆ | ☆ | - | ☆ | - | 4 | 50 |
| Dawson A, et al 2015 | - | - | - | ☆☆ | ☆☆ | ☆ | ☆ | - | 6 | 60 |
| Sims M, et al 2016 | ☆ | - | - | ☆☆ | ☆☆ | ☆ | ☆ | - | 7 | 70 |
| Nadimpalli S, et al 2017 | ☆ | - | - | ☆☆ | ☆☆ | ☆ | ☆ | - | 7 | 70 |
| Simons R, et al 2020† | ☆ | ☆ | - | - | - | - | ☆ | ☆ | 4 | 50 |
| ***Eating behavior*** | | | | | | | | | | |
| Kohlmaier J. 2004 | - | - | - | ☆☆ | ☆☆ | ☆ | ☆ | - | 6 | 60 |
| Harrington E, et al 2006 | - | - | - | ☆☆ | ☆ | ☆ | ☆ | - | 4 | 40 |
| Connolly M, 2011 | ☆ | ☆ | ☆ | ☆☆ | ☆☆ | ☆ | ☆ | - | 9 | 90 |
| Clark J & Winterowd C, 2012 | ☆ | - | - | ☆☆ | ☆☆ | ☆ | ☆ | - | 7 | 70 |
| Hayman L, 2012 | ☆ | ☆ | ☆ | ☆☆ | - | ☆ | ☆ | - | 7 | 70 |
| Johnson P, et al 2012 | - | ☆ | - | ☆☆ | ☆☆ | - | ☆ | - | 6 | 60 |
| Kelly N, et al 2018 | ☆ | ☆ | - | ☆☆ | ☆☆ | ☆ | ☆ | - | 8 | 80 |
| Hoggard L., et al 2019 | - | - | - | ☆☆ | ☆☆ | ☆ | ☆ | - | 6 | 60 |

| | | | | | | | | | | |
|---|---|---|---|---|---|---|---|---|---|---|
| Longmire-Avital B & McQueen C, 2019 | ☆ | ☆ | ☆ | ☆☆ | ☆☆ | ☆ | ☆ | - | 9 | 90 |
| Kelly N, et al 2020 | ☆ | - | - | ☆☆ | ☆☆ | ☆ | ☆ | - | 7 | 70 |
| Hagiwara N, et al 2020 | ☆ | - | - | ☆☆ | ☆☆ | ☆ | ☆ | - | 7 | 70 |
| † *cohort studies* | | | | | | | | | Average | 66% |
